# How the COVID-19 pandemic affects transgender health care in upper-middle-income and high-income countries – A worldwide, cross-sectional survey

**DOI:** 10.1101/2020.12.23.20248794

**Authors:** Andreas Koehler, Joz Motmans, Leo Mulió Alvarez, David Azul, Karen Badalyan, Koray Basar, Cecilia Dhejne, Dragana Duišin, Bartosz Grabski, Aurore Dufrasne, Natasa Jokic-Begic, Antonio Prunas, Christina Richards, Kirill Sabir, Jaimie Vaele, Timo Ole Nieder

**Affiliations:** University Medical Center Hamburg-Eppendorf, Institute for Sex Research, Sexual Medicine, and Forensic Psychiatry, Martinistrasse 52, 20246 Hamburg, Germany; Transgender Infopunt, Ghent University Hospital, Ghent, Belgium; Transgender Europe, Berlin, Germany; La Trobe University, La Trobe Rural Health School, Department of Community and Allied Health, Discipline of Speech Pathology, Melbourne, Australia; Eurasian Key Populations Coalition, Warszawa, Poland; Hacettepe University, Department of Psychiatry, Ankara, Turkey; Karolinska University Hospital, Andrology, Sexual Medicine, Transgender Medicine, Stockholm, Sweden; Clinical Centre of Serbia, Department of Psychiatry, Belgrade, Serbia; Jagiellonian University Medical College, Sexology Lab, Department of Psychiatry, Krakow, Poland; Genres Pluriels, Brussels, Belgium; University of Zagreb, Clinical and Health Psychology Unit, Department of Psychology, Faculty of Humanities and Social Sciences, Zagreb, Croatia; University of Milan, Department of Psychology, Milan, Italy; Tavistock and Portman NHS Foundation Trust & Regents University London, School of Psychotherapy and Psychology, London, United Kingdom; FtM Phoenix Group, Moscow, Russia; University of Waikato, Faculty Arts & Social Sciences, New Zealand

## Abstract

**Background:** Since the beginning of the COVID-19 pandemic, access to medical care was restricted for nearly all non-acute medical conditions. Due to their status as a vulnerable social group and the inherent need for transition-related treatments (e.g., hormone treatment), transgender people are assumed to be affected particularly severely by the restrictions caused by the COVID-19 pandemic. This study aims to assess the impact of the COVID-19 pandemic on the health and health care of transgender people.

**Methods and findings:** As an ad hoc collaboration between researchers, clinicians, and 23 community organizations, we developed a web-based survey. The survey was translated into 26 languages, and participants were recruited via various social media and LGBTIQ-community sources. Recruitment started in May 2020. We assessed demographical data, physical and mental health problems (e.g., chronic physical conditions), risk factors (e.g., smoking), COVID-19 data (symptoms, contact history, knowledge and concerns about COVID-19), and the influence of the COVID-19 pandemic on access to transgender health care and health-related supplies. To identify factors associated with the experience of restrictions to transgender health care, we conducted multivariate logistic regression analysis.

5267 transgender people from 63 higher-middle income and high-income countries participated in the study. Over 50% of the participants had risk factors for a severe course of a COVID-19 infection and were at a high risk of avoiding testing or treatment of a COVID-19 infection due to the fear of mistreatment or discrimination. Access to transgender health care services was restricted due to the COVID-19 pandemic for 50% of the participants. Male sex assigned at birth and a lower monthly income were significant predictors for the experience of restrictions to health care. 35.0% of the participants reported at least one mental health conditions. Every third participant had suicidal thoughts, and 3.2% have attempted suicide since the beginning of the COVID-19 pandemic. A limitation of the study is that we did not analyze data from low-income countries and access to the internet was necessary to participate.

**Conclusions:** Transgender people are assumed to suffer under the severity of the pandemic even more than the general population due to the intersections between their status as a vulnerable social group, their high amount of medical risk factors, and their need for ongoing medical treatment. The COVID-19 pandemic can potentiate these vulnerabilities, add new challenges for transgender individuals, and, therefore, can lead to devastating consequences, like severe physical or mental health issues, self-harming behaviour, and suicidality.

## Introduction

Transgender people experience their gender as incongruent with the sex assigned at birth. They might identify as a binary gender (female, male) or outside of the gender binary. People who are non-binary might experience their gender as moving between male and female (e.g., genderfluid) or as situated beyond the gender binary (e.g., genderqueer). Some reject the concept of sex and gender at all, either on a personal, or a general level (e.g., agender). Transgender health care primarily focusses on medical measures to support the person’s transition to live their gender both physically and socially. This may include hormone therapy, gender-affirming surgery, and a variety of additional interventions (e.g., voice and communication therapy, hair removal)[1]. For treatment-seeking transgender people these interventions positively affect mental health and quality of life, and are thus considered state-of-the-art treatments [2-5]. However, not all transgender people want to undergo any of these types of care, or might have access to it [6, 7].

Up to now, transgender people have been considered to be a vulnerable social group, many of whom have experienced discrimination and marginalization by society, and health care systems in particular [4, 8]. Access to transgender health care is often restricted due to legal requirements, financial barriers, and ‘gatekeeping’ health care providers in countries all over the world [9, 10]. These elements can lead to a negative impact on transgender individual’s health and quality of life [4].

During to the COVID-19 pandemic access to medical care was, and in some areas globally still is, restricted for nearly all non-acute medical conditions. State health care, as well as private practice, were mainly focusing on COVID-19. Since transgender individuals often need access to medical treatments such as hormone therapy [1], these restrictions are likely to have increased psychological distress. Due to both their status as a vulnerable social group and their need for transition-related treatments [4, 8], this impact may be particularly severe for transgender people [11].

Even though some authors have already addressed the impact of the COVID-19 pandemic on transgender people, there remains a dearth of evidence. Perez-Brumer and Silva-Santisteban [12] discussed the disparities transgender people face in Peru due to binary-based policies as a response to the COVID-19 pandemic (only women or men are allowed to leave their homes on certain weekdays) and how this increasing discrimination impacts physical and mental health. Van der Miesen and colleagues [13] summarized the intersections between health care, human rights, and socioeconomic stress for transgender individuals during the COVID-19 pandemic. They state the need for a joint strategy of policy makers, transgender advocates, health care providers, and governments. Wang and colleagues [11] refer to the barriers of care for transgender people in times of COVID-19 in light of evidence from their clinic in Beijing, China. The restricted access to hormone treatment was associated with high levels of depression and anxiety due to the challenge of continuing presenting and socially living as their sex assigned at birth. All the authors state the need for collaborative strategies between relevant stakeholders (e.g., governments, health care providers, advocacy groups) to actively consider the difficulties faced by the transgender population during the COVID-19 pandemic and the need for high-quality evidence to base strategies upon [11-13].

Therefore, as the first of its kind, the current study systematically investigates the impact of the COVID-19 pandemic on the health and health care of transgender individuals. As an ad hoc collaboration between researchers, community members and clinicians from several countries, it aims to generate empirical evidence on the situation of transgender individuals in times of COVID-19 and, therefore, help to develop and implement measures addressing the obstacles that affect transgender individuals during the pandemic.

## Methods

### Study design

The TransCareCovid-19 survey (www.transcarecovid-19.com), is a web-based survey designed to investigate the effects of the COVID-19 pandemic on health care for transgender individuals. This cross-sectional study was developed by XX, YY, and ZZ in cooperation with local health care providers and community members. Several members of the research group are transgender. Additional consultation was sought in the construction of the survey by peer and professional organisations.

The survey was first developed in German and, in cooperation with 23 community organisations (for details see www.transcarecovid-19.com), translated into 27 languages (Arabic, Azerbaijani, Armenian, Bulgarian, Chinese, Croatian, Dutch, English, Farsi, French, Georgian, German, Hungarian, Italian, Kazakh, Kyrgyz, Macedonian, Polish, Portuguese, Romanian, Russian, Serbian, Spanish, Swedish, Tajik, Turkish, Ukrainian).

The study followed strict ethical guidelines and received ethical approval from both the Local Psychological Ethics Committee at the University Medical Center MM-NN (No.: LPEK-0130, date: 01/04/2020), as well as from PP University Hospital (BC-07607, date: 15/04/2020). All respondents provided informed consent.

### Participants

To recognize the heterogeneity of the transgender population, the survey was open to anyone who identifies, experiences, and/or describes themselves, as transgender and were at least 16 years of age. Participants were recruited via postings on LGBTIQ-related social media channels, mailing lists of support groups and LGBTIQ-related associations, and through snowball sampling.

### Data collection

The data collection started in May 2020 and is still ongoing. The participant recruitment was supported by several scientific and community organisation (for details see www.transcarecovid-19.com). The present sample is based on the data collected until August 9, 2020. We choose this date as the COVID-19 pandemic was slowing down in most countries all over the world and our data allowed us to give a comprehensive overview on how the first wave of COVID-19 affected transgender people. This paper consists of analyses of data from high-income and upper middle-income economies according to the World Bank country classification[14] only, as participant recruitment for lower middle-income and low-income economies started later.

### Variables

The TransCareCovid-19 survey collects demographical data regarding age, education, occupational status, country of residence, place of residence, residence status, living situation, financial income, relationship, minority status (person of colour, religious minority, sexual minority, gender minority, minority due to disability status, another minority), sex assigned at birth, and gender. The participants’ country of residence was classified by the current World Bank country classification by income level [14]. Upper middle-income economies are defined as having a gross national income (GNI) per capita of $4,046 – $12,535. A country with a GNI per capita of greater than $12,535 is considered a high-income economy. Physical health problems were assessed using items based on established studies[15, 16] and free-text responses. COVID-19 symptoms, contact history, satisfaction with information, knowledge, and concerns about COVID-19, were assessed following Wang and colleagues using 4-point Likert scales (1-highest value, 4-lowest value) [17]. We added additional items related to transgender-specific discrimination and avoidance of health care based on former studies [16]. Finally, data regarding transition-related health care such as medical procedures which the respondent has undergone, and the influence of the COVID-19 pandemic on access to transgender health care and health-related supplies (e.g., chest binders), were assessed based on an established protocol [18]. Depending on the procedures which the respondent had undergone, participants were specifically asked about the influence of the COVID-19 pandemic on single treatments. For example, only participants who already used hormones where asked if access to their medication was restricted. Fears about future restrictions on treatment were investigated if participants had already sought or planned the treatment.

Due to the method of participant recruitment, access to a web-enabled device, social media activity, and technical affinity need to be considered as potential biases. By encouraging participants to promote the survey with their peers we tried to address the issue of lacking social media activity. Unfortunately, we were not able to provide a paper-pencil version of the survey due to the wide range of the study and a lack of financial and human resources.

### Statistical analysis

Continuous data are presented as mean (SD). Categorical data are presented as n (%). Missing data were deleted pairwise. To identify factors associated with the experience of restrictions to transgender health care, we conducted a multivariate logistic regression analysis. Experiencing restrictions in at least one domain (hormone treatment, hair removal treatment, surgery, surgical aftercare) was included as depended variable (yes, no). We entered the covariates in two blocks. Block 1 included age, education (no formal education, primary, secondary, tertiary education), relationship (yes, no), minority status (person of colour, religious minority, sexual minority, gender minority), disability (yes, no), sex assigned at birth (female, male), and gender (binary, non-binary). This block was labelled Demographic Characteristics. Block 2 included the following aspects of the social environment: Countries’ GNI per capita (upper middle-income economy, high-income economy), residence status (yes, no), population of place of residence (urban, rural), ability to make ends meet with monthly income (very easily, easily, fairly easily, with some difficulty, with difficulty, with great difficulty), and health insurance (yes, no). This block was labelled Social Environment. Cox and Snell’s R 2 and Nagelkerke’s R 2 are reported as coefficients of determination. To check for violations of assumptions for logistic regression such as independence of errors, incomplete information from the predictors, diagnostics were conducted on all relevant variables. Collinearity diagnostics did not reveal significant multicollinearity concerns for any of the variables in the model. A p value of <.05 was considered to be statistically significant. SPSS 24.0 was used for all statistical analyses.

## Results

Between May 1, 2020, and August 9, 2020, 7905 potential participants accessed the survey. 597 participants did not give informed consent and thus were not able to access the survey. Another 593 participants gave informed consent but did not answer any further questions. From the remaining 6715 participants, 1223 individuals were excluded from the analyses because they did not respond to at least 50% of the survey. For the present analysis, 225 participants from low-income and lower middle-income economies were not included. The final dataset consisted of 5267 participants.

On average, the present sample is of early middle age (30.70 years ± 12.06), highly educated (60.7% tertiary education), mostly single (48.7%), and living in an urban environment (79.9%). 57.3% were assigned female at birth. 74.0% identified as binary [trans] man or [trans] woman, whereas 21.5% identified as a non-binary gender.

Basic and transgender-related demographics are presented in Table 1. Table 2 and Fig 1 show the current country of residence of the participants.[14] Most participants lived in European countries. Fig 2 gives an overview of the physical health problems of the participants. 2768 (52.6%) reported a least one acute or chronic condition. 525 (10.0%) reported that these conditions resulted in severe limitations in daily activities, 1674 (31.8%) reported limitations to some extent. 509 (9.7%) had no limitations due to their chronic condition. 1009 (19.2%) of the participants were smokers. 327 (6.2%) used to smoke but had recently stopped. 3035 (73.6%) had at least once in their life seriously considered suicide, 1827 (35.1%) had had suicidal thoughts since the beginning of the COVID-19 pandemic. 1797 (34.5%) had had at least once suicide attempt, 168 (3.2%) have attempted suicide since the beginning of the COVID-19 pandemic.

**Table 1.**
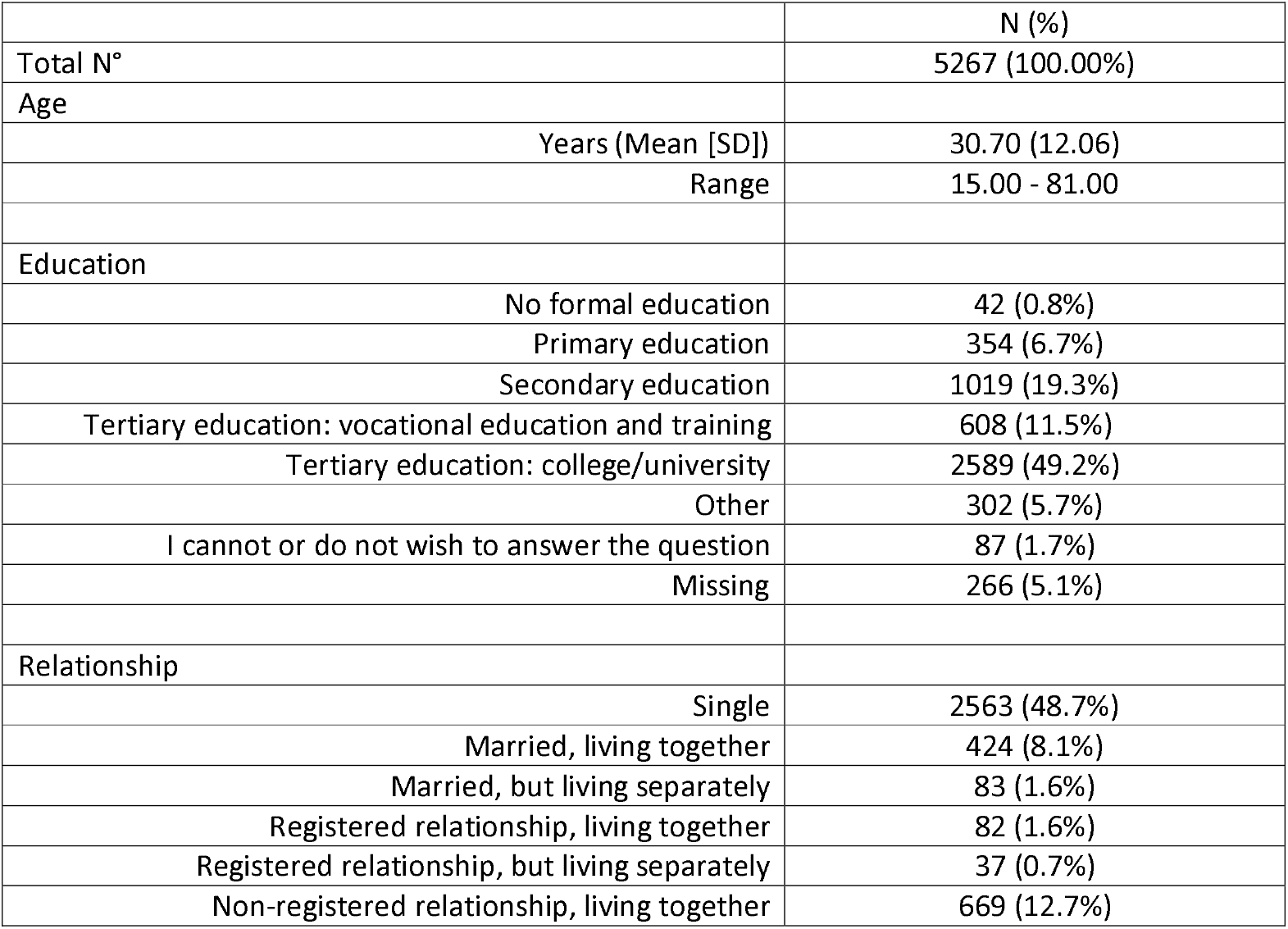
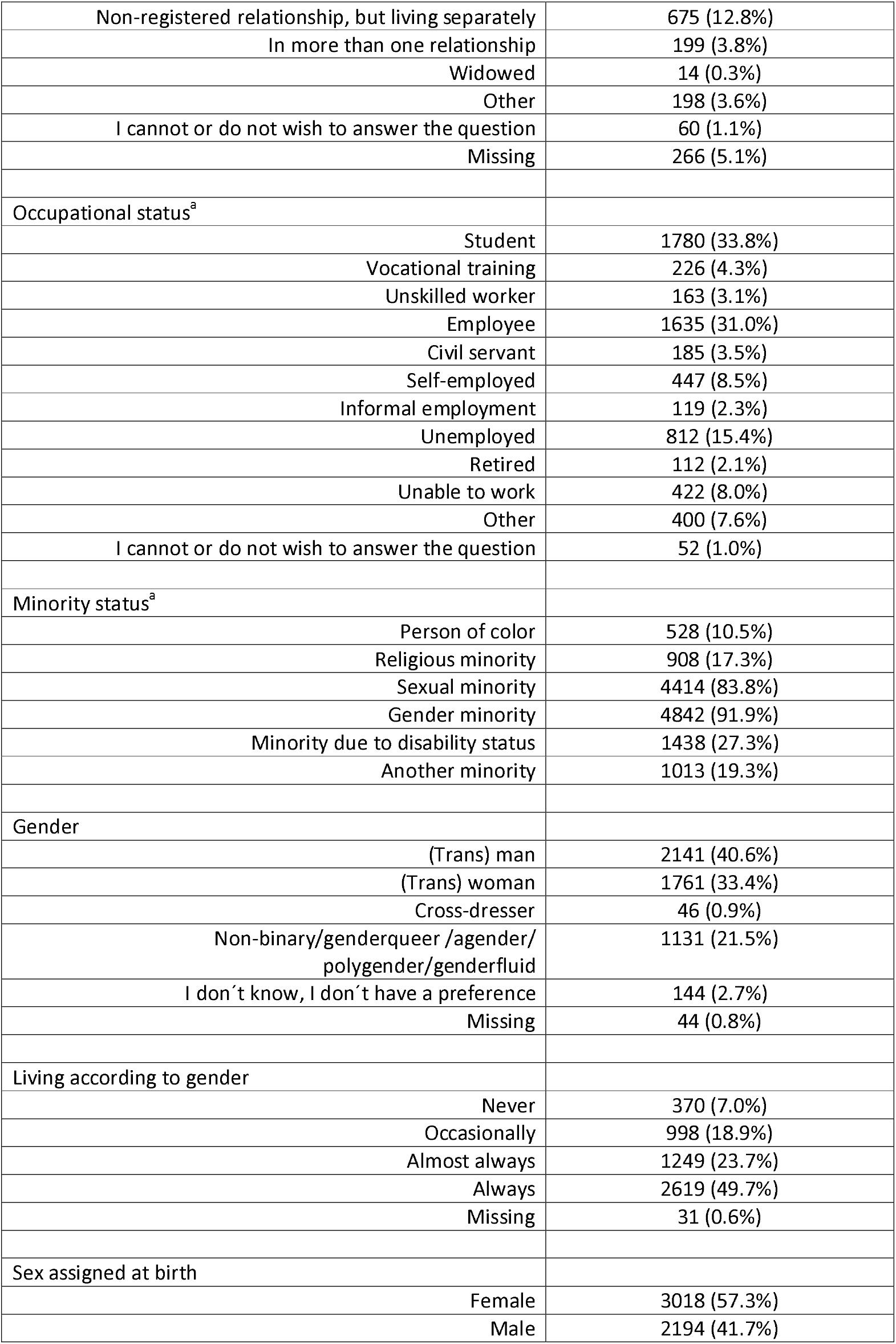
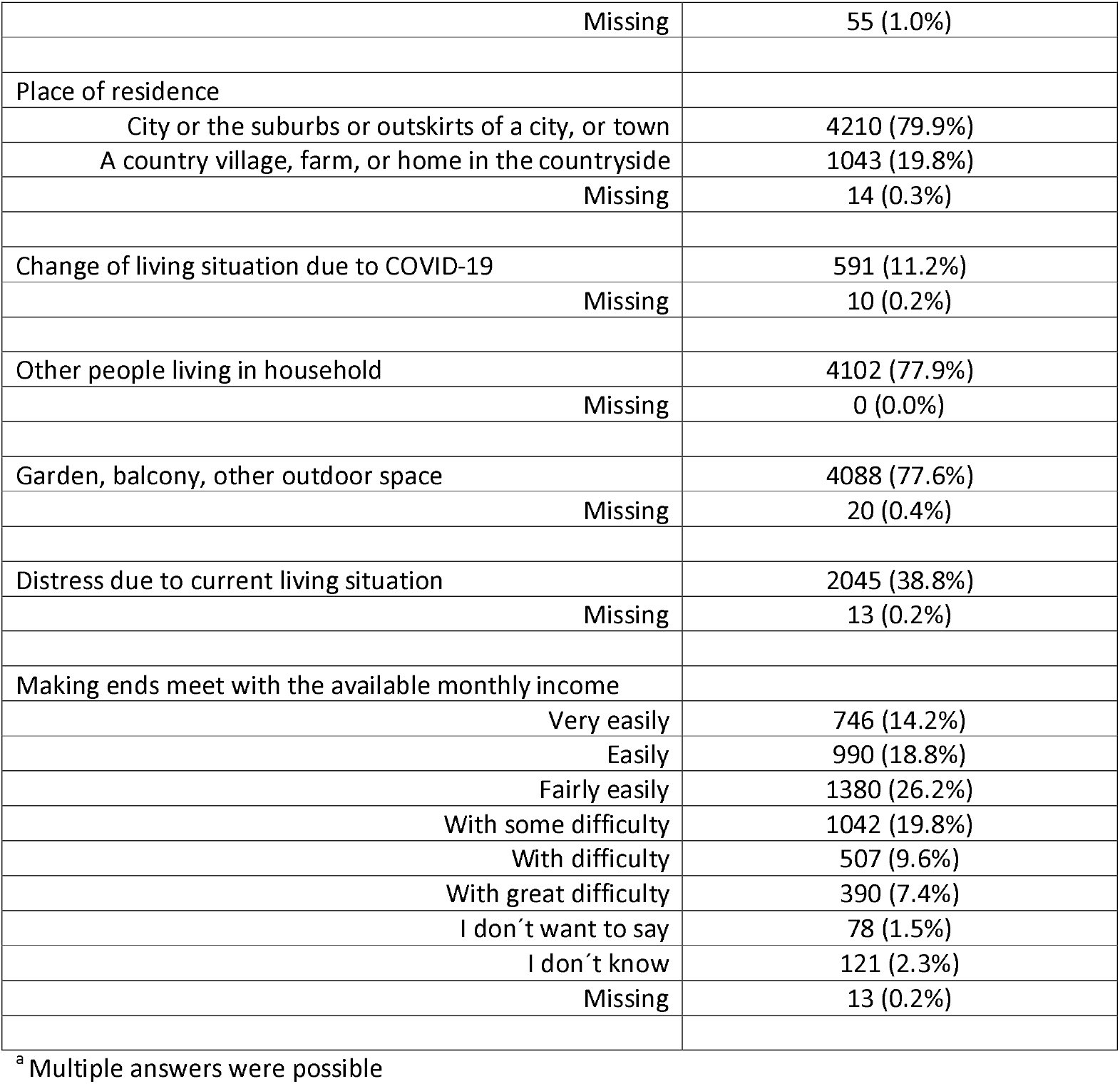
Demographics and social situation

|  | N (%) |
| --- | --- |
| Total N° | 5267 (100.00%) |
| Age |  |
| Years (Mean [SD]) | 30.70 (12.06) |
| Range | 15.00 - 81.00 |
| Education |  |
| No formal education | 42 (0.8%) |
| Primary education | 354 (6.7%) |
| Secondary education | 1019 (19.3%) |
| Tertiary education: vocational education and training | 608 (11.5%) |
| Tertiary education: college/university | 2589 (49.2%) |
| Other | 302 (5.7%) |
| I cannot or do not wish to answer the question | 87 (1.7%) |
| Missing | 266 (5.1%) |
| Relationship |  |
| Single | 2563 (48.7%) |
| Married, living together | 424 (8.1%) |
| Married, but living separately | 83 (1.6%) |
| Registered relationship, living together | 82 (1.6%) |
| Registered relationship, but living separately | 37 (0.7%) |
| Non-registered relationship, living together | 669 (12.7%) |
| Non-registered relationship, but living separately | 675 (12.8%) |
| In more than one relationship | 199 (3.8%) |
| Widowed | 14 (0.3%) |
| Other | 198 (3.6%) |
| I cannot or do not wish to answer the question | 60 (1.1%) |
| Missing | 266 (5.1%) |
| Occupational status<sup>a</sup> |  |
| Student | 1780 (33.8%) |
| Vocational training | 226 (4.3%) |
| Unskilled worker | 163 (3.1%) |
| Employee | 1635 (31.0%) |
| Civil servant | 185 (3.5%) |
| Self-employed | 447 (8.5%) |
| Informal employment | 119 (2.3%) |
| Unemployed | 812 (15.4%) |
| Retired | 112 (2.1%) |
| Unable to work | 422 (8.0%) |
| Other | 400 (7.6%) |
| I cannot or do not wish to answer the question | 52 (1.0%) |
| Minority status<sup>a</sup> |  |
| Person of color | 528 (10.5%) |
| Religious minority | 908 (17.3%) |
| Sexual minority | 4414 (83.8%) |
| Gender minority | 4842 (91.9%) |
| Minority due to disability status | 1438 (27.3%) |
| Another minority | 1013 (19.3%) |
| Gender |  |
| (Trans) man | 2141 (40.6%) |
| (Trans) woman | 1761 (33.4%) |
| Cross-dresser | 46 (0.9%) |
| Non-binary/genderqueer /agender/<br>polygender/genderfluid | 1131 (21.5%) |
| I don't know, I don't have a preference | 144 (2.7%) |
| Missing | 44 (0.8%) |
| Living according to gender |  |
| Never | 370 (7.0%) |
| Occasionally | 998 (18.9%) |
| Almost always | 1249 (23.7%) |
| Always | 2619 (49.7%) |
| Missing | 31 (0.6%) |
| Sex assigned at birth |  |
| Female | 3018 (57.3%) |
| Male | 2194 (41.7%) |
| Missing | 55 (1.0%) |
| Place of residence |  |
| City or the suburbs or outskirts of a city, or town | 4210 (79.9%) |
| A country village, farm, or home in the countryside | 1043 (19.8%) |
| Missing | 14 (0.3%) |
| Change of living situation due to COVID-19 | 591 (11.2%) |
| Missing | 10 (0.2%) |
| Other people living in household | 4102 (77.9%) |
| Missing | 0 (0.0%) |
| Garden, balcony, other outdoor space | 4088 (77.6%) |
| Missing | 20 (0.4%) |
| Distress due to current living situation | 2045 (38.8%) |
| Missing | 13 (0.2%) |
| Making ends meet with the available monthly income |  |
| Very easily | 746 (14.2%) |
| Easily | 990 (18.8%) |
| Fairly easily | 1380 (26.2%) |
| With some difficulty | 1042 (19.8%) |
| With difficulty | 507 (9.6%) |
| With great difficulty | 390 (7.4%) |
| I don't want to say | 78 (1.5%) |
| I don't know | 121 (2.3%) |
| Missing | 13 (0.2%) |

<sup>a</sup> Multiple answers were possible

**Table 2.** Countries

|  |  |
| --- | --- |
| Upper middle-income countries | 762 (14.5%) |
| Argentina | 6 (0.1%) |
| Armenia | 34 (0.6%) |
| Azerbaijan | 19 (0.3%) |
| Belarus | 3 (0.1%) |
| Bosnia and Herzegovina | 3 (0.1%) |
| Botswana | 3 (0.1%) |
| Brazil | 36 (0.7%) |
| Bulgaria | 14 (0.3%) |
| China | 150 (2.7%) |
| Georgia | 27 (0.5%) |
| Indonesia | 2 (0.1%) |
| Iran | 1 (0.1%) |
| Iraq | 1 (0.1%) |
| Kazakhstan | 30 (0.5%) |
| Lebanon | 6 (0.1%) |
| Malaysia | 2 (0.1%) |
| Mexico | 2 (0.1%) |
| Montenegro | 16 (0.3%) |
| Namibia | 4 (0.1%) |
| North Macedonia | 17 (0.3%) |
| Russia | 61 (1.1%) |
| Serbia | 69 (1.3%) |
| South Africa | 75 (1.4%) |
| Turkey | 181 (3.3%) |
| High-income countries | 4505 (85.5%) |
| Australia | 109 (2.0%) |
| Austria | 94 (1.7%) |
| Barbados | 1 (0.1%) |
| Belgium | 229 (4.2%) |
| Canada | 128 (2.3%) |
| Chile | 1 (0.1%) |
| Croatia | 30 (0.5%) |
| Czech Republic | 33 (0.6%) |
| Denmark | 17 (0.3%) |
| Finland | 62 (1.1%) |
| France | 66 (1.2%) |
| Germany | 1311 (23.9%) |
| Greece | 6 (0.1%) |
| Hungary | 74 (1.3%) |
| Iceland | 2 (0.1%) |
| Ireland | 8 (0.1%) |
| Israel | 4 (0.1%) |
| Italy | 48 (0.9%) |
| Japan | 3 (0.1%) |
| Latvia | 1 (0.1%) |
| Liechtenstein | 1 (0.1%) |
| Luxembourg | 2 (0.1%) |
| Malta | 7 (0.1%) |
| Netherlands | 175 (3.2%) |
| New Zealand | 37 (0.7%) |
| Norway | 33 (0.6%) |
| Poland | 296 (5.4%) |
| Portugal | 14 (0.3%) |
| Republic of Korea | 1 (0.1%) |
| Romania | 1 (0.1%) |
| Seychelles | 2 (0.1%) |
| Slovakia | 3 (0.1%) |
| Slovenia | 6 (0.1%) |
| Spain | 136 (2.5%) |
| Sweden | 328 (6.0%) |
| Switzerland | 303 (5.5%) |
| United Kingdom | 563 (10.3%) |
| United States | 366 (6.7%) |
| Taiwan | 1 (0.1%) |
| Missing | 38 (0.7%) |

**Fig 1.**
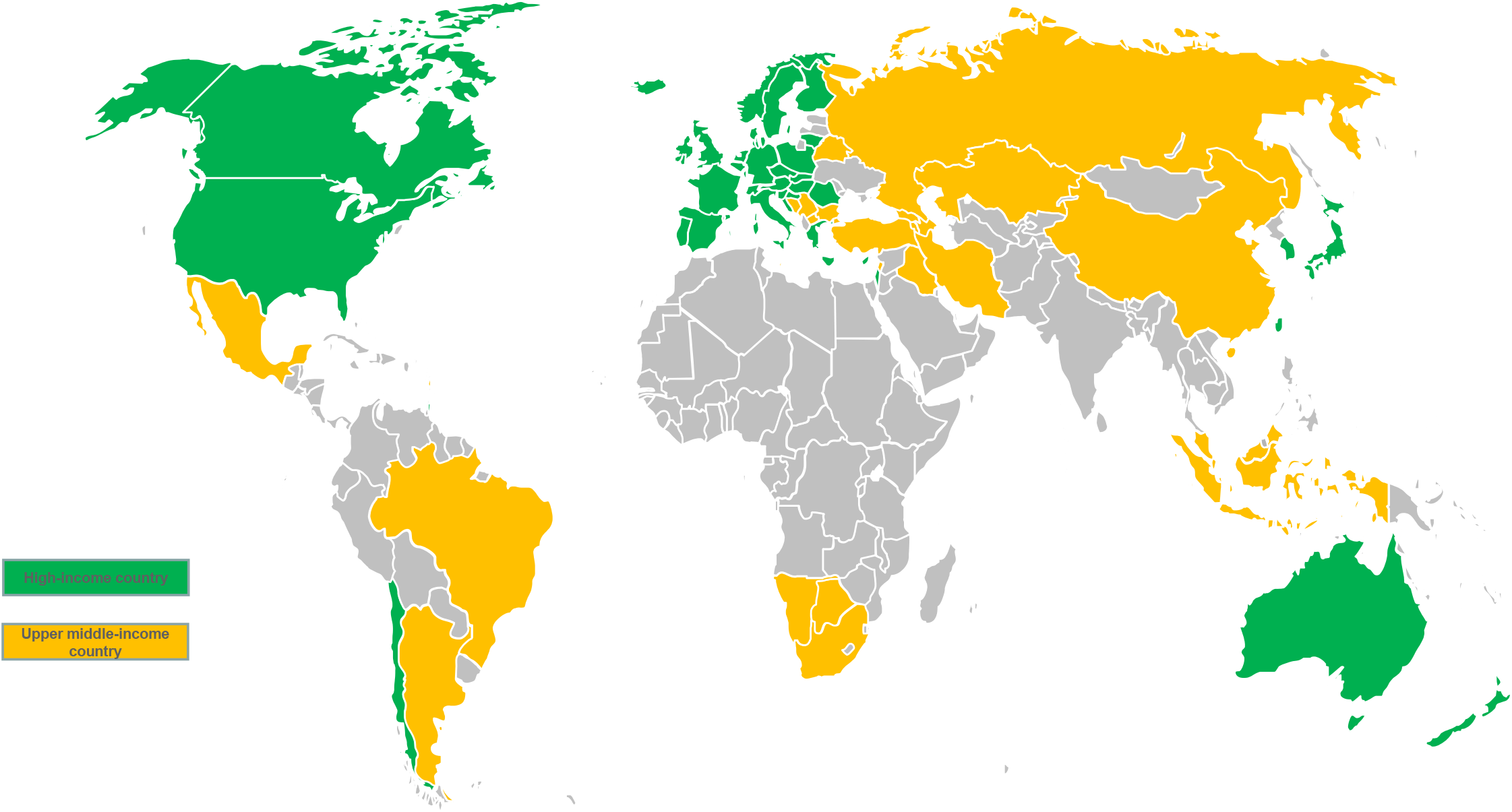
Participants current country of residence

**Fig 2.**
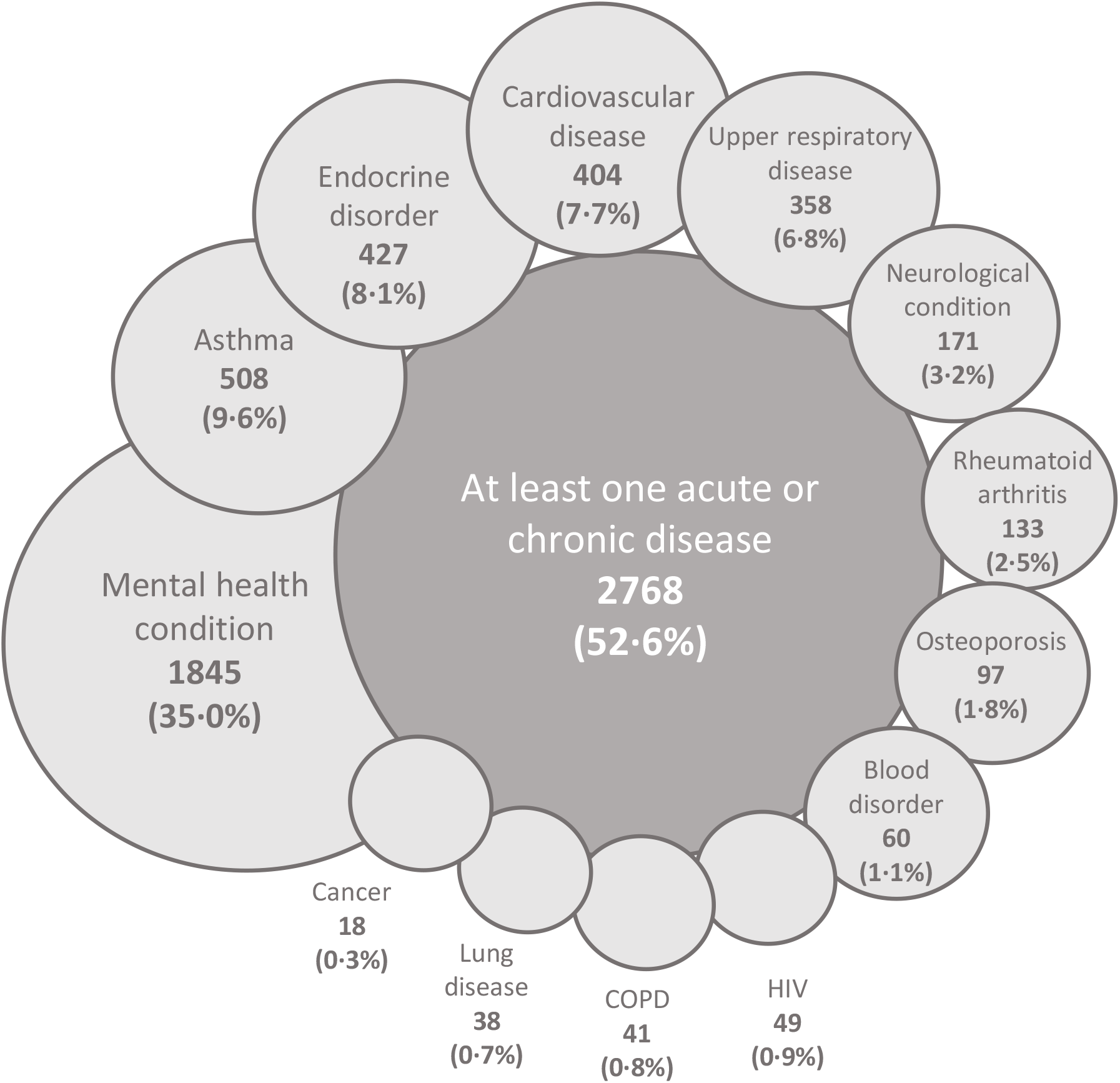
Acute and chronic conditions associated with a risk of a severe course of a COVID-19 infection

Table 3 gives an overview on COVID-19-related topics. Participants were quite satisfied with the information they received on the COVID-19 pandemic (M: 2.13 ± 0.8) and had high confidence in medical personnel to diagnose COVID-19 (M: 2.01 ± 0.8). Participants assumed it as not very likely to be diagnosed with COVID-19 (M: 2.90 ± 0.9) and as highly likely to survive a COVID-19 infection (M: 1.63 ± 0.8). They were concerned that members of their families or close friends will contract COVID-19 (M: 2.09 ± 0.9).

**Table 3.** COVID-19 health care, avoidance, and knowledge

|  |  |
| --- | --- |
| COVID-19 health care |  |
| Tested for COVID-19 within the last 14 days | 130 (2.5%) |
| In quarantine within the last 14 days | 1481 (28.1%) |
| Diagnosed with COVID-19 | 20 (0.4%) |
| Recovered from COVID-19 | 14 (0.3%) |
| Direct contact with a person diagnosed with COVID-19 | 109 (2.1%) |
| Indirect contact with a person diagnosed with COVID-19 | 245 (4.7%) |
| Contact with a person suspected to have COVID-19 | 818 (15.5%) |
| COVID-19 health care avoidance |  |
| Avoided COVID-19 testing because of fear of mistreatment | 295 (5.6%) |
| Avoided COVID-19 testing because of fear of discrimination | 356 (6.8%) |
| Will avoid COVID-19 testing because of fear of mistreatment | 761 (14.4%) |
| Will avoid COVID-19 testing because of fear of discrimination | 888 (16.9%) |
| COVID-19 knowledge |  |
| Transmission via droplet infection | 5040 (95.7%) |
| Increasing number of infections worldwide | 5091 (96.7%) |
| Increasing number of deaths worldwide | 5067 (96.2%) |
| Increasing number of survivals worldwide | 4592 (87.2%) |

Table 4 summarises the influence of the COVID-19 pandemic on transgender health care. Of the 4699 participants that had undergone transition-related treatment or planned to do so, 2875 (61.2%) feared access restrictions to transgender health care in the future due to the COVID-19 pandemic. Of the 3463 participants that had already undergone transition-related treatment, 1706 (49.3%) experienced restrictions in access to transgender health care services. A multivariate logistic regression analysis revealed that the sex assigned at birth and the adequacy of the monthly income predicted the experience of restrictions to transgender health care significantly. Participants assigned male at birth (OR: 2.239) and individuals with a lower income (OR: 1.102) were at greater risk of experiencing restrictions to transgender health care procedures (Table 5).

**Table 4.** Restrictions regarding transgender health care due to the COVID-19 pandemic

|  | N (%<sup>a</sup>) |
| --- | --- |
| Accessed at least one transgender health care procedure | 3463 (65.7%) |
| Experienced restrictions in access to transgender health care | 1706 (49.3%) |
| Accessed or planned at least one transgender-related treatment procedure | 4699 (89.2%) |
| Fear restrictions to transgender health care in the future | 2875 (61.2%) |
| Access to hormones currently restricted | 676 (21.8%) |
| I cannot get a prescription | 162 (24.0%) |
| I cannot get an appointment with my hormone prescriber | 249 (36.8%) |
| A scheduled appointment was canceled without replacement | 110 (16.3%) |
| An appointment has been postponed | 81 (12.0%) |
| I am afraid to go to a medical provider or a hospital | 189 (28.0%) |
| My hormones cannot be supplied or can only be supplied at a lower dosage than has been prescribed to me | 177 (26.2%) |
| Other | 224 (33.1%) |
| Fear that access to hormones will be affected in the future | 2316 (44.0%) |
| Access to hair removal treatment currently restricted | 665 (63.4%) |
| Fear that access to hair removal treatment will be affected in the future | 776 (44.1%) |
| Surgery cancelled or postponed |  |
| Yes | 454 (15.6%) |
| Not yet, but I expect it will | 568 (19.5%) |
| Problems with aftercare for recent surgery | 344 (56.4%) |
| I cannot get an appointment for aftercare | 45 (13.1%) |
| A scheduled appointment was canceled without replacement | 70 (20.3%) |
| An appointment has been postponed | 84 (24.4%) |
| Complications (e.g., secondary bleeding) have not been treated | 22 (6.4%) |
| I am afraid to go to a doctor or a hospital | 41 (11.9%) |
| Other | 82 (23.8%) |
| Fear that surgical aftercare will be affected in the future | 316 (21.9%) |
| Access restricted to |  |
| medical material necessary after surgery (e.g., vaginal dilators) | 160 (3.0%) |
| other material (e.g., binders, packing material) | 651 (12.4%) |
| non-medical supplies (e.g., wigs, make-up) | 860 (16.3%) |
| Access to counselling services (e.g., peer counselling) limited | 2285 (43.4%) |
| Alternative options for accessing counselling services | 1271 (47.9%) |
| Undergoing mental health care (e.g., for depression) | 2055 (39.0%) |
| Access to mental health care limited | 1271 (61.9%) |
| Fear that access to mental health care will be affected in the future | 1035 (50.4%) |
| Member of a support group | 1543 (28.5%) |
| Access to support groups limited | 1067 (69.2%) |

<sup>a</sup> Percentages refer to the number of participants who assessed a certain transgender health service, not to the total sample

**Table 5.** Multiple regression analysis regarding the experience of restrictions to transgender health care

|  | b ± SE | CI | p | OR (95% CI) |
| --- | --- | --- | --- | --- |
| Demographics |  |  |  |  |
| Age | 0.000 ± 0.003 | -0.007 - 0.006 | 0.899 | 1.000 (0.993, 1.006) |
| Education | -0.013 ± 0.067 | -0.148 - 0.119 | 0.850 | 0.987 (0.869, 1.121) |
| Relationship | 0.033 ± 0.079 | -0.123 - 0.201 | 0.677 | 1.033 (0.886, 1.205) |
| POC | 0.158 ± 0.145 | -0.129 - 0.436 | 0.269 | 1.171 (0.885, 1.550) |
| Religious minority | -0.083 ± 0.118 | -0.320 - 0.144 | 0.466 | 0.920 (0.742, 1.142) |
| Sexual minority | 0.000 ± 0.116 | -0.220 - 0.236 | 0.997 | 1.215 (0.891, 1.657) |
| Gender minority | 0.194 ± 0.157 | -0.104 - 0.490 | 0.217 | 1.081 (0.891, 1.657) |
| Disability | 0.077 ± 0.095 | -0.097 - 0.276 | 0.388 | 0.380 (0.903, 1.657) |
| Sex assigned at birth | 0.967 ± 0.082 | 0.809 - 1.136 | 0.001 | 2.239 (2.239, 3.092) |
| Non-binary gender | -0.034 ± 0.119 | -0.262 - 0.213 | 0.767 | 0.966 (0.769, 1.214) |
| Social Environment |  |  |  |  |
| Countries GNI per capita | -0.167 ± 0.115 | -0.392 - 0.056 | 0.153 | 0.846 (0.677, 1.057) |
| Residence status | -0.312 ± 0.254 | -0.845 - 0.182 | 0.212 | 0.732 (0.449, 1.191) |
| Population place of residence | 0.106 ± 0.102 | -0.092 - 0.320 | 0.292 | 1.111 (0.913, 1.354) |
| Monthly income | 0.097 ± 0.028 | 0.045 - 0.150 | 0.003 | 1.102 (1.043, 1.164) |
| Health insurance | 0.093 ± 0.175 | -0.255 - 0.422 | 0.600 | 1.098 (0.792, 1.521) |
| Constant | 0.754 ± 0.554 | -0.283 - 1.840 | 0.172 | 0.808 |

R<sup>2</sup> 0.063 (Cox & Snell), 0.084 (Nagelkerke). Model $\chi^2$ (15) = 186.693, p=0.000

## Discussion

As the first of its kind, the current study provides empirical insights into how the COVID-19 pandemic affects the health care of transgender people in higher middle-income and high-income countries. Over 50% of our participants had risk factors for a severe course of a COVID-19 infection and were at a high risk of avoiding testing or treatment of a potential COVID-19 infection due to fear of mistreatment and/or discrimination. Transgender health care was highly affected by the COVID-19 pandemic, with almost half of participants experiencing some sort of restricted access to health care. Restricted access to transgender health care applied rather universally to all transgender individuals, with a male sex assigned at birth and a lower monthly income as the only significant predictors.

We found, that over 50% of transgender individuals have risk factors for a severe course of a COVID-19 infection (i.e., pre-existing medical conditions, being a smoker).[19-22] Compared with that, 23.5% of the working population in the EU and one-third of the European general population over 15 suffer from chronic conditions [23]. Similar numbers were found for other OECD countries [24]. Only in the US is the prevalence of chronic conditions between the transgender and the general population approximately equal, with 6 out of 10 adults having a chronic condition [25]. Additionally, our participants were likely to avoid COVID-19 testing or care because of fears of discrimination or mistreatment. This was true even though they were aware of the potential severity of COVID-19. This avoidance of care could even worsen their risk for serious consequences of COVID-19.

We found that access to all transgender health care interventions was restricted due to the COVID-19 pandemic. This could be especially important for those who access ongoing treatments such as hormone therapy [3]. Hormone therapy is considered one of the most important treatment options for many transgender individuals and is highly associated with better mental health and quality of life [26]. An interruption of a hormone therapy is not associated with physical risks when gonadectomy was not performed. However, mental health risks must be considered. Stopping hormone treatment is often associated with a return of some features related to the sex assigned at birth (e.g., the return of menstruation). Moreover, mood swings and symptoms of depression and anxiety can occur [2, 3, 27]. There is a risk of psychological distress for the twenty percent of our sample whose hormones have been, or still are, restricted due to the pandemic. This association has also been reported by Wang and colleagues in their clinic in Beijing, China [11]. The same may well be true for hair removal treatments. While hair removal might not have the same value as hormone treatment, it is often performed as a supportive measure as hormone treatment alone could be insufficient to eliminate hair growth [27]. As a population, transgender individuals are already experiencing poor mental health, decreased quality of life, and are at high risk of discrimination and harassment [4, 8]; consequently, the restrictions in access to health care may affect transgender people to a greater extent than the general population. This higher vulnerability might also intersect with other minority statuses, such as being a person of colour.

All these transgender-specific issues caused by the COVID-19 pandemic need to be addressed by both structural changes as well as counselling services (e.g., peer counselling) and mental health care professionals. However, due to the COVID-19 pandemic, access to these measures is limited, too. For 43.4% of the participants who assessed counselling services, the access to those was restricted. 61.9% of those who undergo mental health care (e.g., for a pre-existing affective disorder) experienced access restrictions. Only half of the participants had alternative options for accessing counselling services (e.g., online consultations). Additionally, even access to low-threshold services, like support groups, was limited for two thirds of their members. In light of the available data, the COVID-19 pandemic and its associated restrictions, combined with the systemic inequalities transgender people face in almost every country in the world, appears to have had a significant impact on the (mental) health of transgender people. Tragically, our study found that more than one third of our sample had had suicidal thoughts and 3.2% had attempted suicide since the beginning of the pandemic. And even though no causal conclusions can be drawn based on our cross-sectional study, it seems reasonable to assume that many of these suicidal attempts are linked to the situation caused by the COVID-19 pandemic. Indeed, at the least this association ought to be a warning sign that transgender people might be disproportionally affected by COVID-19.

Some treatment options for transgender people such as genital surgeries [1] have been highly restricted due to the COVID-19 pandemic, too. At the time of the data analysis, approximately 15.6% of surgeries were already cancelled and another 19.5% of the participants expected their surgery to be cancelled. Additionally, aftercare for recent surgery was also restricted. As genital surgery is considered an important treatment option to increase quality of life and reduce psychological distress in transgender people [2, 28], a postponement of these procedures can negatively impact a person’s well-being. Especially regarding surgical aftercare, restrictions could lead to serious health concerns like wound infections or disorders of wound healing [2]. Moreover, due to insecurities caused by the postponement of surgery, mental health problems could occur which should be addressed by counselling services or mental health professionals when possible. However, as the access to these services is restricted too, problems associated with postponed surgeries might not be addressed sufficiently and could intersect with already existing distress such as restrictions to accessing hormone treatment.

All potential distress caused by restrictions to transgender health care might be further affected by social and socio-economic circumstances, as already reported by Perez-Brumer and Silva-Santisteban [12]. We found that one third of our participants had difficulties making ends meet with their available monthly income and another third experienced distress due to their current living situation. However, using regression analysis, we only found a male sex assigned at birth and a lower monthly income to be significant predictors for the experience of restrictions to transgender health care. The significant association to a male sex assigned at birth could be due to a greater stigma towards transgender people assigned male at birth [29]. The association between the monthly income and a higher risk of restrictions to transgender health care with an OR of 1.102 could be considered weak. Therefore, it seems that the COVID-19 pandemic hits the transgender population in higher-middle-income and high-income countries in its entirety and that there aren’t any major protective social factors.

The strength of our study is that it includes a high number of participants from several countries all over the world. This is important as, with a number of 6.8 to 355 per 100,000, transgender people are only a small group within the general population [30]. The results are therefore likely to be applicable to other upper middle-income and high-income countries with similar health systems. Moreover, the study was developed and conducted by transgender researchers in close cooperation with community organisations. Therefore, we intentionally took power differences between researchers and study subjects into account and tried to ensure tangible outcomes for the community.

We consider the type of data collection a potential limitation of our study due to access to the internet being a necessity. This could have excluded participants with low income or without experience with digital technology. However, our sample was comparable to previous studies, in terms of gender or education, for example [7, 18]. As there is still a dynamic COVID-19 situation, with increasing and decreasing numbers of infections and changing national measures to address these, this research also did not explore international comparisons. In countries where COVID-19 has been more effectively suppressed the impacts of COVID-19 could, therefore, be lower. Moreover, we only analysed data from higher middle-income and high-income countries, which is why the results cannot be generalized to countries with lower GNI per capita. However, as participant recruitment in lower middle-income countries and low-income countries appeared to be difficult (but is still ongoing) the clear need for evidence of how COVID-19 affects transgender people led us to focus on the present data. We currently strive to provide data from lower middle-income countries and low-income countries as soon as possible.

In sum, it appears that the COVID-19 pandemic has an extraordinary impact on the transgender population in higher middle-income and high-income countries, and that transgender people might suffer under the severity of the pandemic even more than the general population. This is due to the intersections between their status as a vulnerable social group, their high amount of medical risk factors, and their need for ongoing medical treatment. The COVID-19 pandemic can potentiate these vulnerabilities, add new challenges for transgender people, and can therefore lead to devastating consequences such as severe physical or mental health issues, self-harming behaviour, and even suicidality.

## Supporting information

STROBE statement

## Data Availability

We will consider sharing de-identified, individual participant-level data that underlie the results reported in this Article on receipt of a request detailing the study hypothesis and statistical analysis plan. All requests should be sent to the corresponding author. The corresponding author and lead investigators of this study will discuss all requests and make decisions about whether data sharing is appropriate based on the scientific rigour of the proposal. All applicants will be asked to sign a data access agreement.

## Acknowledgements

A worldwide survey needs a lot of helping hands. Annette Güldenring (Heide, Germany and Bundesverband Trans*) as well as Petra Weitzel and Lena Balk (both dgti, Germany) actively supported us in the development of the questionnaire in the original version in German. Julia Kata from Trans-Fuzja was also very helpful in distributing the questionnaire in Poland. The data from Mainland China would not have been possible without the help of Eddy Xu and the team of The Trans Well-Being Project. For the USA, Asa Radix, MD, and Ayden Scheim, PhD, were also very helpful.

## Contributors

XX, YY, and ZZ conceived the study. All authors reviewed the study concerning country-specific issues. All authors contributed to participant recruitment and data collection. XX, YY, and ZZ managed the data and did the statistical analysis. All authors collaborated in interpretation of the results and drafting and revision of the manuscript.

