## Supplementary material for "How the COVID-19 pandemic affects transgender health care in upper-middle-income and high-income countries – A worldwide, cross-sectional survey": STROBE statement

STROBE Statement—Checklist of items that should be included in reports of ***cross-sectional studies***

|  | Item No | Recommendation |
| --- | --- | --- |
| **Title and abstract** | 1 | (*a*) Indicate the study’s design with a commonly used term in the title or the abstract  *The title describes the study design as “a worldwide, cross-sectional study”.* |
|  |  | (*b*) Provide in the abstract an informative and balanced summary of what was done and what was found  *The abstract describes the methods and findings.* |
| Introduction | | |
| Background/rationale | 2 | Explain the scientific background and rationale for the investigation being reported  *The background and rationale are described in the Introduction, paragraphs 2 & 3.* |
| Objectives | 3 | State specific objectives, including any prespecified hypotheses  *The specific aims of the study are stated in the Introduction, paragraphs 4.* |
| Methods | | |
| Study design | 4 | Present key elements of study design early in the paper  *The study design is discussed in the subsection “Study design” of the Methods section.* |
| Setting | 5 | Describe the setting, locations, and relevant dates, including periods of recruitment, exposure, follow-up, and data collection  *The institutional setting and study locations are described in the subsection “Study design” of the Method section.* |
| Participants | 6 | (*a*) Give the eligibility criteria, and the sources and methods of selection of participants  *Selection of the sample is discussed in the subsection “Participants” of the Methods section.* |
| Variables | 7 | Clearly define all outcomes, exposures, predictors, potential confounders, and effect modifiers. Give diagnostic criteria, if applicable  *All variables assessed in the study are discussed in the “Variables” subsection.* |
| Data sources/ measurement | 8* | For each variable of interest, give sources of data and details of methods of assessment (measurement). Describe comparability of assessment methods if there is more than one group  *Measurement of the variables are discussed in the “Variables” subsection.* |
| Bias | 9 | Describe any efforts to address potential sources of bias  *Potential bias is discussed in the last paragraph of the “Variables” subsection.* |
| Study size | 10 | Explain how the study size was arrived at  *Sample determination is discussed in the ”Participants” and “Data collection” subsection of the Methods section.* |
| Quantitative variables | 11 | Explain how quantitative variables were handled in the analyses. If applicable, describe which groupings were chosen and why  *Use of variables is discussed in the Statistical Analysis subsection.* |
| Statistical methods | 12 | (*a*) Describe all statistical methods, including those used to control for confounding  *Statistical methods are discussed in the “Statistical Analysis” subsection.* |
|  |  | (*b*) Describe any methods used to examine subgroups and interactions  *Described in the “Statistical analysis” subsection.* |
|  |  | (*c*) Explain how missing data were addressed  Missing observations are discussed in the “*Statistical analysis” subsection.* |
|  |  | (*d*) If applicable, describe analytical methods taking account of sampling strategy  *None.* |
|  |  | (*e*) Describe any sensitivity analyses  *None.* |
| Results | | |
| Participants | 13* | (a) Report numbers of individuals at each stage of study—eg numbers potentially eligible, examined for eligibility, confirmed eligible, included in the study, completing follow-up, and analysed  *Data collection completion rates are discussed in the Results section.* |
|  |  | (b) Give reasons for non-participation at each stage  *Discussed in the Results section.* |
|  |  | (c) Consider use of a flow diagram  *No STROBE flowchart was prepared, as the inclusion and exclusion of participants was already discussed in detail within the first paragraph of the Results section.* |
| Descriptive data | 14* | (a) Give characteristics of study participants (eg demographic, clinical, social) and information on exposures and potential confounders  *Participants characteristics are presented in Table 1.* |
|  |  | (b) Indicate number of participants with missing data for each variable of interest  *Discussed in the Results section.* |
| Outcome data | 15* | Report numbers of outcome events or summary measures  *Both numbers and percentages/proportions are reported throughout the Results*  *Section.* |
| Main results | 16 | (*a*) Give unadjusted estimates and, if applicable, confounder-adjusted estimates and their precision (eg, 95% confidence interval). Make clear which confounders were adjusted for and why they were included  *Unadjusted results are presented for all outcomes.* |
|  |  | (*b*) Report category boundaries when continuous variables were categorized  *Not applicable.* |
|  |  | (*c*) If relevant, consider translating estimates of relative risk into absolute risk for a meaningful time period  *Not applicable.* |
| Other analyses | 17 | Report other analyses done—eg analyses of subgroups and interactions, and sensitivity analyses  *Not applicable.* |
| Discussion | | |
| Key results | 18 | Summarise key results with reference to study objectives  *Results are summarized in paragraph 1 of the Discussion section.* |
| Limitations | 19 | Discuss limitations of the study, taking into account sources of potential bias or imprecision. Discuss both direction and magnitude of any potential bias  *Limitations are discussed in paragraph 5 of the Discussion section* |
| Interpretation | 20 | Give a cautious overall interpretation of results considering objectives, limitations, multiplicity of analyses, results from similar studies, and other relevant evidence  *Final paragraph of Discussion section.* |
| Generalisability | 21 | Discuss the generalisability (external validity) of the study results  *The representativeness of the sample is discussed in paragraph 5 of the Discussion section.* |
| Other information | | |
| Funding | 22 | Give the source of funding and the role of the funders for the present study and, if applicable, for the original study on which the present article is based  *“Role of the funding source” subsection in the Methods section.* |

*Give information separately for exposed and unexposed groups.

**Note:** An Explanation and Elaboration article discusses each checklist item and gives methodological background and published examples of transparent reporting. The STROBE checklist is best used in conjunction with this article (freely available on the Web sites of PLoS Medicine at http://www.plosmedicine.org/, Annals of Internal Medicine at http://www.annals.org/, and Epidemiology at http://www.epidem.com/). Information on the STROBE Initiative is available at www.strobe-statement.org.
